# A study on the appropriate use of topdown approach for stepping up economic activities in districts of different States/Union Territories in India

**DOI:** 10.1101/2020.06.11.20091322

**Authors:** Arindom Chakraborty, Kalyan Das

## Abstract

At the beginning of phase 4.0 of lockdown, every State /Union Territories(UT) needs to take appropriate mitigation efforts (lockdown and testing) that may change red zones to orange and even to green zones by May 31, 2020. On the contrary, negligence in following the guidelines (for interventions) strictly, may be alarming and may change green zone status to even the worst red zone status. This has been established through a Statistical model based study here. From the present investigation, the Government can decide the right measures to take up so as to reduce the transmission of the virus and to open partial economic activities in different districts of a state. The whole idea can also be extended to containment zones in a district with sufficient data at hand.

## 1. Introduction

Even though lockdown has been extended to the fourth round, districts (cities) in some states in India are still surging ahead with the number of cases. A long period of complete nationwide lockdown (shutting off all trade, construction and other activities) is in fact causing tremendous economic setback. Despite huge economic package has recently been declared for weaker sections and MSME (Micro, Small and Medium enterprises), it is too critical to cope with, unless economic activities are at least partially resumed in some regions where the situation is much better. Keeping that in mind the Government of India (GOI) has made a master plan by introducing colour coded zones where red, orange and green zones indicate the degree of severity in the spread of infection in decreasing order. But the problem is that due to high spread character of the corona virus, there may be a chance that a green zone with no sign of infection at any day may subsequently develop some cases (Orange or red zone) over a period of time due to free movement(relaxation of lockdown). This particular work, through a mathematical model, is basically recommending the government to act accordingly by observing the change in the colour code.

The policy taken by the Government to follow the topdown approach to resist the downfall of economy is certainly a thoughtful and effective one. This approach essentially indicates a tradeoff between a systematic control of the outbreak and the economic activity. The idea is to manage the Covid-19 pandemic at micro level by identifying all districts across states of the country into red, orange and green zones and then focusing on to containment zones of any red/orange district. This will enable gradual increment of economic activities adhering to all guidelines in non red zones even during the extended period of the nationwide lockdown. In fact, the third phase of nationwide lockdown with partial relaxations kicked-in on May 01, is a sort of experiment done to fulfill this target of stepping up economy. The lockdown relaxations in various states are essentially based on the incidence of Covid-19.

The nationwide lockdown in India has just stepped into the fourth phase. But is it worthwhile to go for complete lockdown throughout or partial relaxation is enough in some districts of a state where situation is under control ? As noted by economists, the containment of the coronavirus outbreak through extending the mandatory stay-at-home period to 54 days from 40 days in third phase results in a direct output loss of more than 10% over India’s economy. Battered by the fallout of the lockdown that is now in its eighth week, many states are in favour of resuming economic activities outside containment zones. Obviously, the districts and its subzones where economic activities could be resumed adhering to all guidelines, need to be cautiously selected in order to check the spread.

The health and family welfare ministry used two criteria to classify the districts as red zones — the absolute number of cases (at least 6 cases) and the speed of growth in cases. At the begining of 3rd phase of lockdown on 4th May, the ministry has identified 170 red zone districts, 207 orange zone districts reporting cases and 359 green zone districts not reporting any cases across the country. These numbers will change with the fresh cases of novel coronavirus infection. Any red zone district with more than 15 cases would be treated as a district witnessing outbreak and more strict outbreak containment measures would be taken in that zone.The orange colour will be earmarked for those districts where there are less than 15 cases of COVID-19 and there has been no increase in the number of positive cases. In such areas, limited activities like opening of a few public modes of transport, harvesting of farm products will be allowed. In green zones all economic activities are permitted except opening of schools, colleges, restaurants, large gatherings and malls. Certainly it becomes questionable whether any district in a state which has been currently declared as a red zone (an orange zone) would go up (down) to the next upper (lower) zone in subsequent period of time Alternatively, it may certainly be of interest to the government to find out the level of possible interventions that may be adopted for changing the colour code of a zone (district). As there is currently no vaccine or specific drug for COVID-19, the only way, the transmission can be mitigated and controlled, is through good hygiene, isolating suspected cases, and by social distancing measures such as cancelling large events and closing schools. Amongst many other interventions, extension of lockdown beyond 17th May and also aggressive testing are most effectively the two by which the isolating suspected cases, and social distancing can be managed.

Some of the most contested questions regarding the pandemic revolve around testing whether India has been able to ramp-up enough testing and states with low rates of testing paying a higher price in terms of higher covid-related fatalities.

All out efforts are underway as hundreds of thousands of testing kits have become available, and more testing companies and laboratories have been approved. Testing needs to be expanded exponentially as well as strategically as a tool to provide epidemiological evidence.

The purpose of the current investigation is to recommend the appropriate decision on lockdown levels along with enhanced testing, in order to push up the red /orange zone to the upper zone over a period of time. We propose a mathematical model that helps us to go for Bayesian prediction of a colour zone when enhanced testing is done and appropriate lockdown status is followed. Our analysis suggests that with enhanced testing, Government might be able to ease lockdown restrictions more or earlier. This may be due to the reason that, people are perhaps more aware and, hence, more confident about the spread of covid-19 in their respective districts.

## 2. Methods

There have been substantial literature on the statistical prediction of the spread and mortality due to COVID19, eversince the outbreak occurred in Wuhan, China. Linton, et al.(Jan.26, 2020)[1] consider the estimation of incubation period for this corona virus infection. Kiesha Prem and his colleagues (March25, 2020) [2] and Ferguson et.al. (March16,2020) [3] observe the effect nonpharmaceutical interventions for reducing the mortality and infected cases.Tidman (March19,2020) [4] considers a case study to see the effect of blanket testing on the new incidents of infection.

With a small number of infected individuals in a large population, exponential growth models seem to match the reality. But it fails to act as a good model once a large number of people have been infected. As then, gradually the chance of an infected person contacting a susceptible person declines with a growing fraction of people either recovers or develops some level of immunity. Eventually, the chances of an infected person contacting a susceptible person becomes low enough so that the rate of infection decreases. This will lead to fewer cases and eventually at one time, will stop the viral spread.

In this investigation, rather than using an exponential growth model, we consider a model where infection is dependent on the previous days’ incidence scaled by the reproduction number. This time varying number is essentially scaled by a factor involving different levels of lockdown (in the sense of restrictions imposed) and testing component. The scales of lockdown are categories of a variable denoted as follows:

- No lockdown (free movement) represented by category 0
- Partial lockdown represented by category 1
- Complete lockdown (as implemented in hotspots) represented by category 3

Under various levels of lockdown and enhanced testing we compute the time varying reproduction number R_t_. In order to predict the future lockdown status to determine the future colour code of any zone based on the current status we consider the empirical distribution of the reproduction number under different levels of lockdown together with enhanced intensive testing and compute for each *j* = 0,1, 2

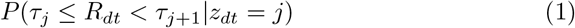

wheres *τ_j_*s are pre-specified cut off points.

The number of infected cases at any day can be predicted using the reproduction number and the weighted average of previous days’ affected figures with the discretized serial interval distribution probability of secondary infection as weights.

## 3. Result

Due to paucity of time, in this investigation, we considered districts from the following states: Maharashtra, Gujarat, Karnataka and Tamil Nadu. However, this district level prediction can be made for other districts from other states. Under the current scenario, i.e. full lockdown and current rate of testing, we considered values at 17th May, 2020 as base. We tried to predict the situation of each of the districts under consideration at 31st May, 2020. It may be noted that this prediction depends on values upto 30th May, 2020 which are once again simulated from the posterior distribution. We found that lockdown still remains the most powerful non-pharmaceutical intervention (NPI),for controlling R_t_, till date for all districts. However, we have already mentioned that this lockdown has direct negative impact on Indian economy. In this situation, in this study, we tried to judge the scenario if the lockdown can partially be lifted for some districts alongwith introducing another non-pharmaceutical intervention (NPI)-intensive testing. If a combination of partial or no lockdown with intensive testing can contain the infection progression, it will be highly beneficial for Indian economy.

To classify any district as red, orange or green, Government of India has given some criteria on the basis of several factors like, time-to-new confirmed case, doubling rate, contribution to total confirmed cases etc. We feel that values of *R_t_* alone can mimic all these behaviors. This motivates us to use a classifier on the basis of simulated *R_t_* values. We classify each district on the basis of empirical probabilities of *R_t_* lying within a pre-specified interval under different NPIs. These intervals are chosen on the distributions of estimated values of *R_t_*.

In this work, for any district *d* at time *t*, we consider the following:

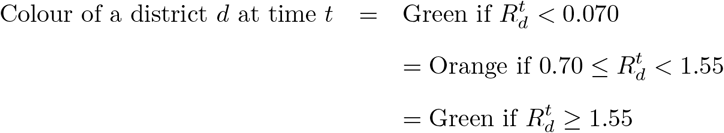

For Karnataka, the results are given in Figure 1a - 1d. The present situation i.e. on 17th May, 2020 is given in figure 1a. Figures 1b-1d predict situations on 31st May, 2020.According to this model, if the lockdown remains present in full force, then without any other NPI, all districts of Karnataka may turn out to be green zones (1b). From 1c, it can be seen that removing lockdown may bring a lot of green districts into threat zones (red or orange) with the current rate of testing. However, an interesting feature for this state can be seen in figure 1d where partial lockdown is assumed with intensive testing. From 1d, the prediction says with the help of intensified testing, these red or orange districts may become green at the end of this month even if partial lockdown is imposed. This clearly indicates that enhanced testing is another powerful NPI alongwith lockdown to combat the spread of COVID-19 in this state.

**Figure 1:**
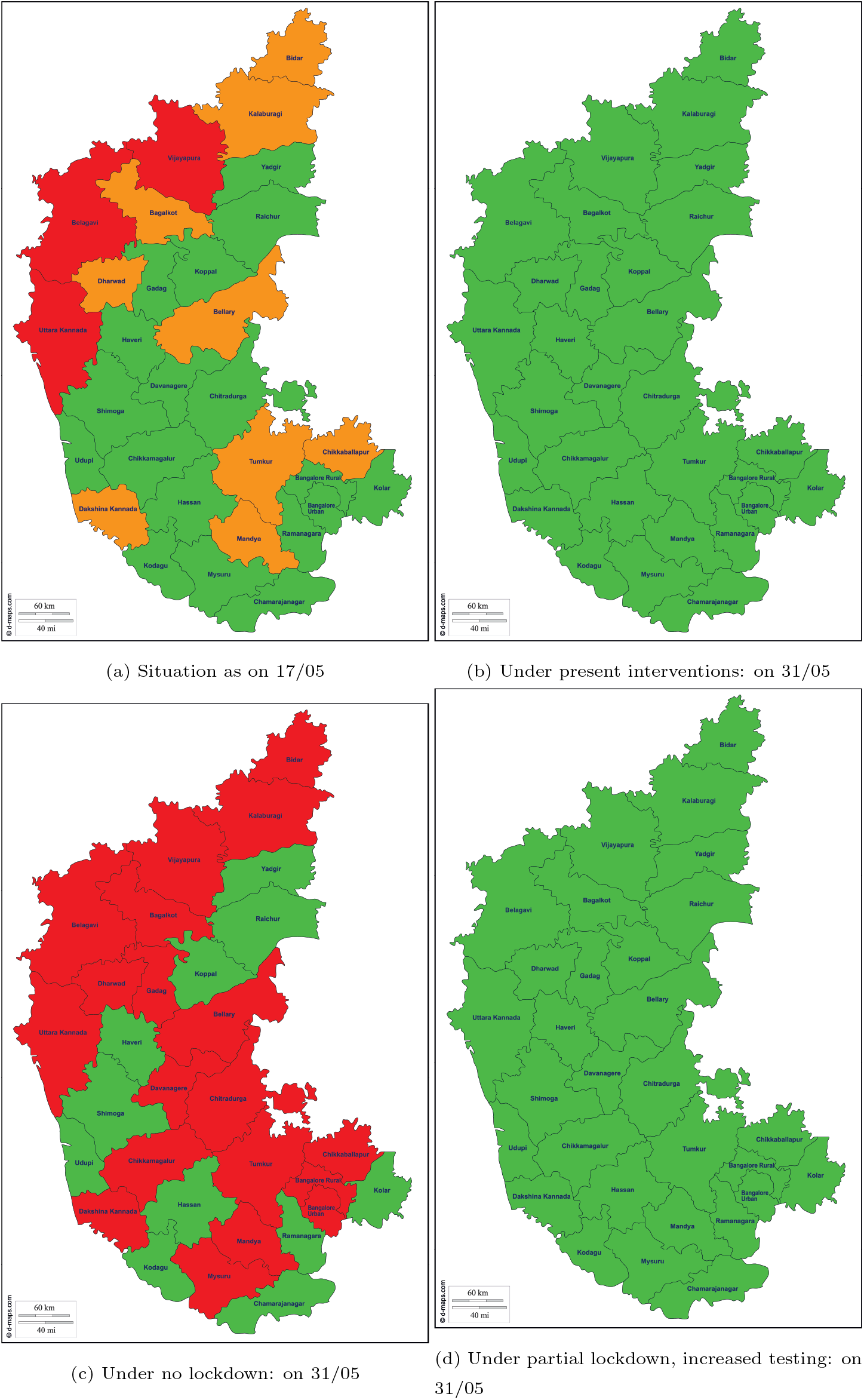
Predicted total new infections for Karnatka under four situations

For Maharashtra, predictions are made under different NPIs and results are given in figures 2a-2d. Like Karnataka, figure 2a represents the current situation of Maharashtra. Figure 2b depicts the situation of the state on 31st May, 2020 under current interventions. It can be seen that Mumbai (both city and suburban) remains in the red zone even if strict measures like full lockdown and intensified testing are implemented. For Mumbai, to get it out from the red zone may require some more time and all NPIs should be maintained with strict adherence. Complete lockdown with present rate of testing may bring a few red districts to one-step-down orange districts (figure 2b). Similar predictions may be made for districts, which are currently in the orange zone,which may turn out to be green ones. As expected, complete withdrawal of lockdown may bring a lot of districts to danger zones. Those who were in the green may turn out to be orange and those districts in orange zones, may find themselves in the red zone at the end of this month (figure 2c). Figure 2d indicates that introduction of partial lockdown and intensified testing may help in restoring the districts into better situations.

**Figure 2:**
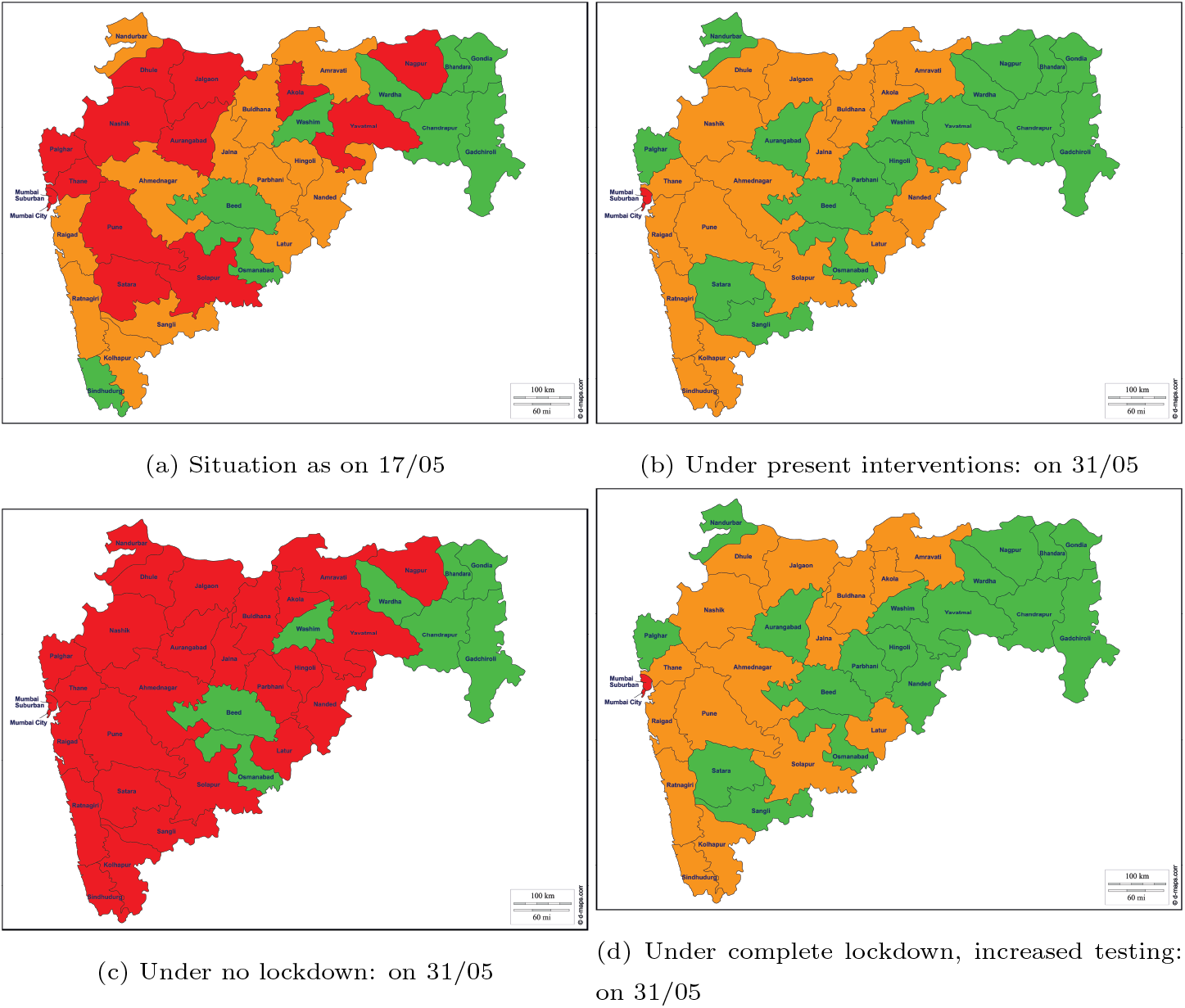
Predicted total new infections for Maharastra under four situations

Among all states, Gujarat has the highest reproduction rate at any given point of time. It can be seen from figures 3a that in the present scenario many of the important districts like Ahmedabad, Surat, Gandhinagar are in red zone. Lifting the complete lockdown will make situation even worse (figure 3c). Interestingly, the intervention like intensive testing is showing most effective results in the presence of complete lockdown for Gujarat (figure 3d). It may be noted from this figure that we have considered complete lockdown, not partial lockdown. Our results show that partial lockdown may not contain the spread even if intensified testing is introduced. We feel that the result is way optimistic as all the districts have turned out to be green under these two NPIs at their highest levels. But we feel that strict adherence of complete lockdown and intensified testing for this state is the need of the hour.

**Figure 3:**
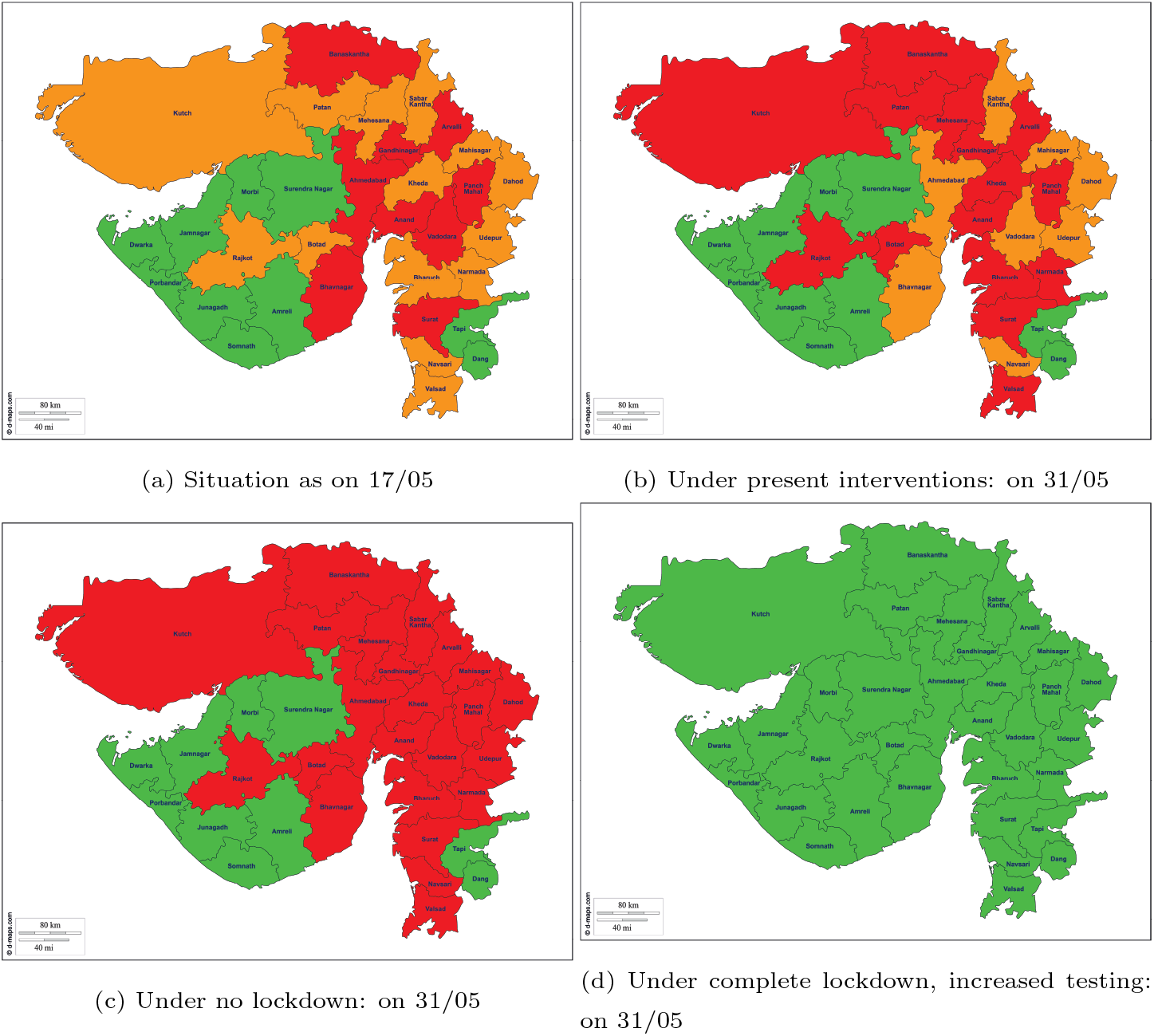
Predicted total new infections for Gujarat under four situations

The situation in Tamil Nadu is becoming very alarming. The spread of this virus was delayed in this state and suddenly it got accelerated in recent times. Figures 4a-4d describe the situation for this southern state. Like Guajarat, here also partial lockdown may not help. Removing lockdown for next two weeks may turn more districts into red ones 4c. Figure 4d indicates that intensive testing may be highly beneficial if it is implemented alonwith highest level of lockdown. In a nutshell we may conclude that imposing complete lockdown for next two weeks may help to curb the spread of this virus for districts which are in orange or green zones. This is found to be true for Maharastra, Gujarat and Tamil Nadu. However, in case of Karnataka, partial lockdown may be imposed.The effect of intensive testing is found to be positive for the districts which are in red zone. But alone this intervention cannot make any point unless complete lockdown is imposed in the severely affected districts.

**Figure 4:**
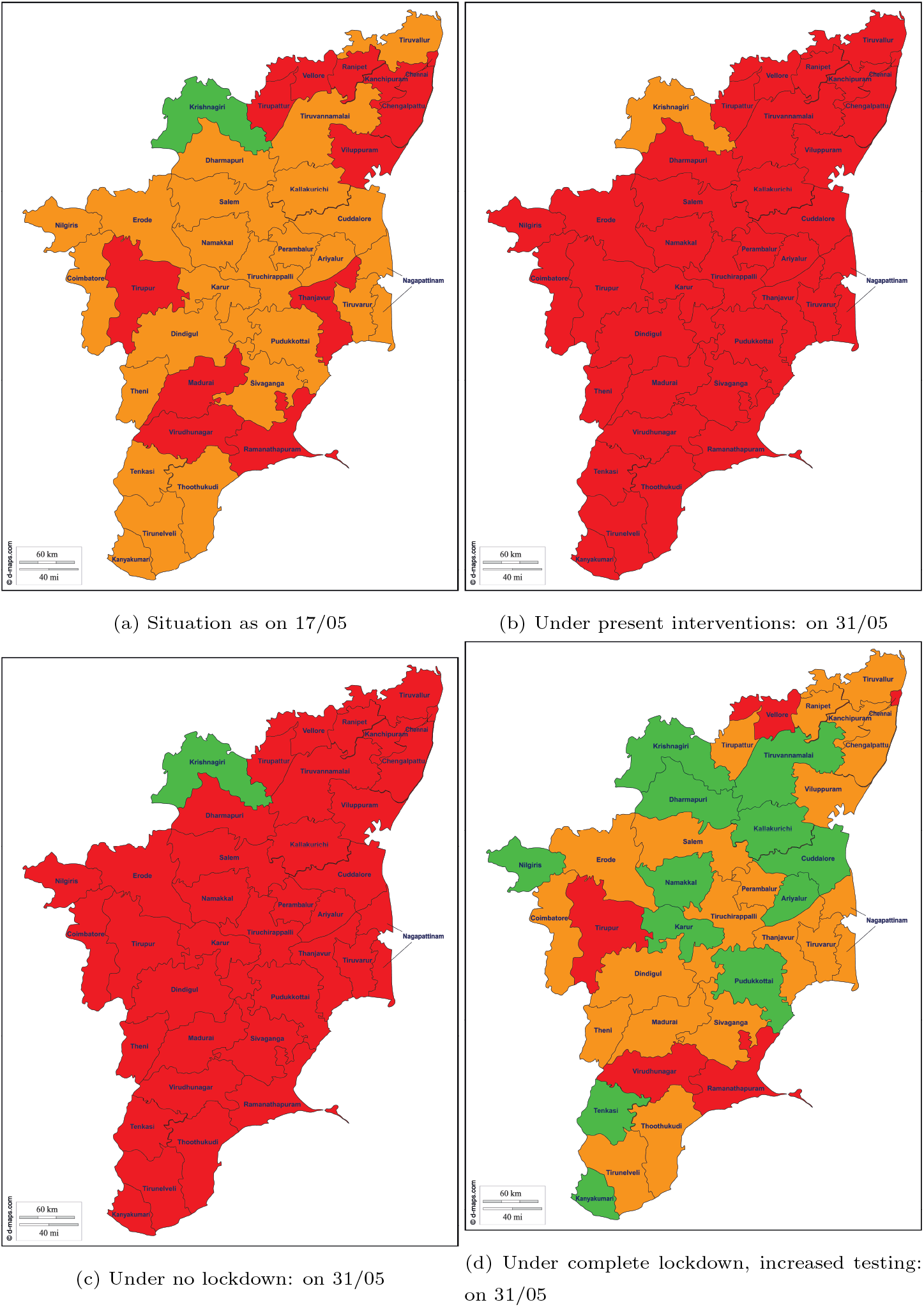
Predicted total new infections for Tamil Nadu under four situations

## 4. Limitations

In this study, the choice of cut-off may require some more consideration. We have not considered any spatial model where the boundary districts may play an important role. The quantification of partial lockdown and intensive testing are policy decisions to be taken by the administration which may be subjective. We also assume that if a person if found to be positive he will be immediately put into isolation.

These predictions are made on the basis of district level data that are available till 9th May, 2020. A lot of total confirmed cases (around 14,000) could not be incorporated due to lack of basic information. For example, this exercise could not be carried out for Delhi as district level data are not in good numbers. However, aggregate data are available on the website. Availability of more data points at district level may change the predictions. This model will be updated as and when more data are at our disposal.

## 5. Discussion

As per the new guidelines issued by Ministry of Home Affairs Government of India for Lockdown 4.0, States/UTs have now been given the power to decide on the delineation of red, green and orange zones. Though States are given more power to take control of the situation going ahead, they will have to make these decisions while abiding by the parameters listed by the Ministry of Health and Family Welfare (MoHFW) and GOI. The new lockdown guidelines essentially pave the way for most economic activity across the country to restart while giving states and union territories more authority on deciding the level of restrictions. In the present study, partial lockdown stands for lockdown based on new guidelines set by GOI.

Self-quarantine and control strategies implemented by government would reduce the effective reproduction number of COVID-19 below one. However, it is necessary to continue social distancing and control travelling to prevent intensity of outbreak. Awareness of the basic reproduction number of corona virus would in fact be useful for calculating vaccination coverage when a safe and effective vaccine against corona virus would come out.

Basic reproduction number is an important parameter which is used to estimate the incidences of COVID-19 outbreak. Technically, the coloured zones in our investigation are selected based on the empirical behavior of the reproduction number. The lockdown levels and aggressive testing play a major role in updating the reproduction number.

In order to prevent health-care system overload in such a pandemic, after the relaxation of lockdown conditions for some zones in a state, a dire necessity is to incorporate testing, contract tracing, and localised Quarantine of suspected cases. Modelling such a strategy (like what proportion of the residents to be tested and how regularly testing could be done given asymptomatic and pre symptomatic transmission) would be extremely useful to guide for controlling the pandemic when lockdown measures are lifted at least partially. In view of the fact that disease dynamics are ultimately greatly influenced by human behaviour, it seems in fact quite natural to have an individual specific component in infectious disease modelling.

## Data Availability

The data is available in public domain.

https://api.covid19india.org/

## 6. Data and software

For this work, we have used the data available in COVID19 Tracker Project (https://api.covid19india.org/) and updated till 9th May, 2020. All computations have been done using RStudio and Stan softwares.

## References

[1] N. M. Linton, T. Kobayashi, Y. Yang, K. Hayashi, A. R. Akhmetzhanov, S. M. Jung, B. Yuan, R. Kinoshita, H. Nishiura, Incubation period and other epidemiological characteristics of 2019 novel coronavirus infections with right truncation: A statistical analysis of publicly available case data, J Clin Med. 9(2) (Feb 17, 2020) 538. doi:doi:10.3390/jcm9020538.

[2] K. Prem, T. W. Liu, and Y. Russell, A. J. Kucharski, N. Eggo, R. M. and Davies, C. for the Mathematical Modelling of Infectious Diseases COVID-19 Working Group, M. Jit, P. Klepac, The effect of control strategies to reduce social mixing on outcomes of the covid-19 epidemic in wuhan, china: a modelling study, Lancet Public Health S2468–2667(20) (2020 Mar 2) 30073-6. doi:doi:10.1016/S2468-2667(20)30073-6.

[3] N. M. Ferguson, D. Laydon, G. Nedjati-Gilani, N. Imai, K. Ainslie, M. Baguelin, S. Bhatia, A. Boonyasiri, A. Z. Ghani, Impact of non-pharmaceutical interventions (npis) to reduce covid19 mortality and healthcare demand, https://mcacs.org/multimedia/files/COVID19.pdf (2020).

[4] Z. Tidman, Coronavirus: Italian village reports no new infections for days after blanket testing, https://www.independent.co.uk/news/world/europe/coronavirus-vo-euganeo-blanket-testing-venetoluca-zaia-a9411201.html (March 19, 2020).

